# Current Estimates of the Economic Burden of Hearing Loss in India: A Societal Cost-of-Illness Study

**DOI:** 10.64898/2026.09.01.26361987

**Authors:** Sunny Mannava, Vidya Ramkumar, G.V.S. Murthy

## Abstract

**Introduction:** Hearing loss (HL) affects over 1·5 billion people globally and India shares a disproportionately high burden including Disabling Hearing Loss (DHL). HL affects an Individual socio-economically, but there are limited studies on the broader societal economic consequences of HL in India.

**Methods:** Using Cost-of-Illness (COI) approach, we studied the societal economic burden of HL in India. This study uses epidemiological and macroeconomic data and modelling to estimate the loss of Gross National Income (GNI) due to HL and DHL across three economic pathways. Uncertainty is evaluated using deterministic and Probabilistic Sensitivity Analyses (PSA).

**Results:** The model estimates that there are in India, 289 million and 85·9 million people with HL and DHL respectively. Direct Loss of GNI and Indirect Loss of GNI (Caregiver burden) are estimated as INR 4,648·4 billion (USD 55·6 billion) and INR 3,268 billion (USD 39 billion) respectively. The Loss of GNI due to Low Education amongst those with HL is estimated as INR 1,041·9 billion (USD 12·45 billion).

**Discussion:** Economic burden of HL is presented across three pathways with Direct Loss of GNI due to DHL being the greatest. It also presents age stratified caregiver economic burden. The findings of the study help in estimating similar cost pathways, advocacy, and policy decisions towards reducing HL prevalence in India and LMICs. This study also highlights the need for India specific estimations related to the HL attributable low education, state-wise disaggregates, and prevalence studies. Funding This study has not received any funding.

## Introduction

An estimated 1·5 billion people globally live with some degree of hearing loss and it has been projected to grow to 2·5 billion by 2050.(1,2) South Asia accounts for 28·2% of the global prevalence of Disabling Hearing Loss (DHL).(2) Given India’s large population and rapidly aging demographic profile, the country is expected to contribute substantially to the future global burden of HL. Each year in India, 23 million babies are born, among whom, 150,000 (0·6%) are born with HL and about 0·1% are born with profound HL.(1,3) HL imposes substantial health and economic burdens globally and in India, where existing data gaps necessitate targeted cost-of-illness analyses.(4)

HL is associated with lower employment rates and reduced income among adults;(5) it also doubles the risk of depression, leads to speech delays, and poor school participation in children.(6) Adults with HL often experience reduced labor force participation, workplace productivity limitations, and communication barriers that may contribute to substantial indirect economic losses.(7,8) Early screening and timely intervention lead to not only health but also economic benefits; a study from South India shows that providing hearing aids averts up to INR 42,000 in disability-adjusted life years (DALYs).(9)

With limited number of audiologists and ENT clinics in India, access remains limited leading to major challenges in scaling screening, diagnosis, and rehabilitation services. Few studies estimated the cost benefits associated with implementing pediatric hearing screening, diagnosis, and rehabilitation using village health workers in a community setting and integrated tele-diagnosis.(10,11) Economic evaluations were undertaken to understand the cost-effectiveness of providing hearing aids to adults.(12) There are partial economic studies estimating the healthcare costs associated with cochlear implants such as the out-of-pocket costs (OOPC) to inform insurance plans.(13) A report published by the World Health Organization (WHO) in 2017 estimated the productivity costs and cost to the education system associated with HL for various countries including India. However, these estimates were based on data extrapolated from other countries and 2015 Global Burden of Disease (GBD) study.(1,14)

Cost-of-Illness (COI) studies help in understanding the economic burden of a disease and provide policy-makers and various healthcare stakeholders a case for advocacy. COI studies also help in understanding the various cost drivers and funding needs related to screening or programmatic costs for a particular disease or a condition as observed from various studies.(15,16) In our literature review, India specific COI studies related to HL were not available, and continue to remain so.(17)

In the current study, we adopted a societal cost-of-illness framework to estimate the economic burden of hearing loss in India using nationally derived demographic, epidemiological, and macroeconomic parameters across multiple economic pathways.

## Methods

### Study Design

The cost-of-illness methodology was used to estimate the economic burden of hearing loss in India using a societal perspective for the year 2024. This prevalence-based cross-sectional study considered population, prevalence parameters, and national-level demographic and economic data for India. The economic burden is estimated as three primary cost pathways using a top-down approach, each reflecting a distinct dimension of economic impact.

1. **Direct Loss (DL) of Gross National Income (GNI)**: Quantifies the costs due to unemployment attributed to disabling hearing loss (DHL) amongst adults.
2. **Indirect Loss of GNI (IL) or Caregiver Cost**: Represents the productivity time lost by family members who provide unpaid care to individuals with HL and DHL.
3. **Loss to the Economy due to Low Educational Attainment (LE)**: Accounts for the income deficit due to reduced school participation and low academic achievement because of HL.

Figure 1 outlines how costs are stratified by age group and categorized into the three pathways to assess national-level welfare losses due to HL. The prevalence data were stratified by age group (<18 years, 18– 60 years, and >60 years). Key macroeconomic indicators, such as employment-to-population ratios, labor force participation, GDP per capita, and GNI, were used to model direct and indirect costs.(18,19) The study used secondary data and where empirical estimates for India were unavailable, proxy values and assumptions informed by global literature were used (e.g., caregiver burden estimates from blindness literature,(20) productivity loss ratios from U.S. data).(21) Discounting was not applicable given the one-year prevalence-based time horizon.

**Figure 1:**
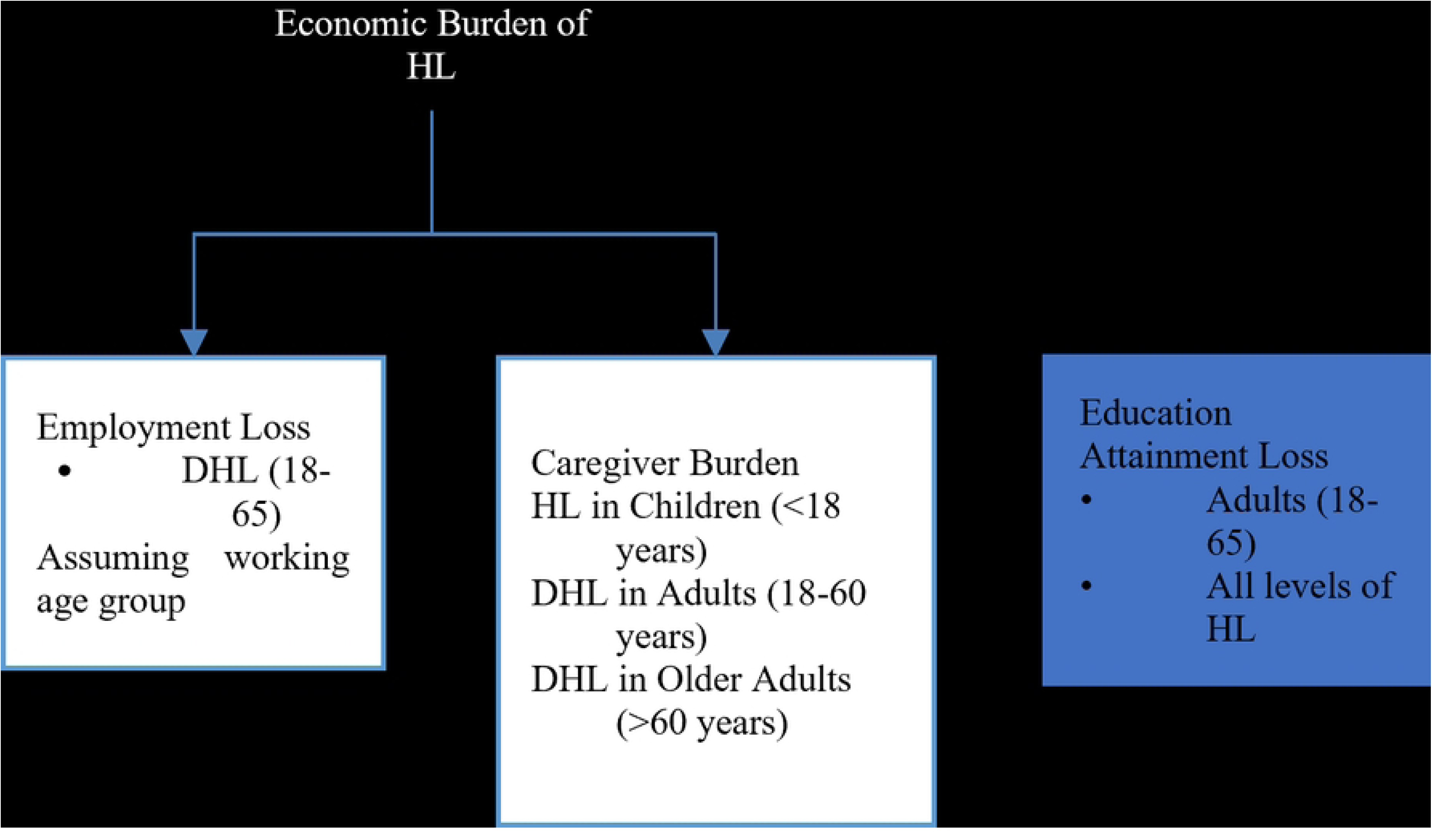
Stratification of the economic burden of hearing loss (HL) in India (2024).

All monetary estimates are expressed in 2024 constant rate Indian rupees (INR), and where relevant, converted to billions for reporting. We have used the International Monetary Fund (IMF) and World Bank deflator to convert INR to International Dollars (Int$) and United States Dollar (USD) respectively. The R statistical environment (v4.4.3) was used for all simulations, data processing, analyses, and graphical visualization. The full set of model equations, symbol definitions, and distribution-fitting procedures is provided in the appendix.

### Definitions

- **Hearing Loss (HL)**: Hearing threshold >25 dB HL in the better hearing ear across all age groups.(1)
- **Disabling Hearing Loss (DHL)**: Hearing threshold >40 dB HL in adults in the better ear.(1)
- **Adults with Low Education:** Individuals aged >18 years whose highest qualification is less than secondary education.(22)

### Population and Prevalence Estimates

Demographic estimates for India’s child and adult populations were derived from the World Bank estimates, published literature, and government reports for the year 2024. Prevalence rates of HL and DHL were applied separately to children (<18 years), working-age adults (18–60 years), and older adults (>60 years). The number of individuals with HL and DHL was calculated by applying prevalence rates to population counts, Table 1 shows the parameters used for the analysis.

**Table 1:** Prevalence and total number of people with HL and DHL in various age groups.

| Age Group | Population | HL<br>Prevalence | DHL<br>Prevalence | HL Count | DHL Count |
| --- | --- | --- | --- | --- | --- |
| <18 years | 420,771,379 | 0·12 | NA* | 48,535,979 | NA* |
| 18–60 years | 877,816,154 | 0·16 | 0·04 | 144,400,757 | 33,080,186 |
| >60 years | 152,348,258 | 0·63 | 0·35 | 96,207,925 | 52,864,846 |
| Total (N) | 1,450,935,791 | - | - | 289,144,661 | 85,945,032 |

\*NA: Not
Applicable.

### Macroeconomic Parameters

Key economic inputs included:

- Per Capita GNI Purchasing Power Parity (PPP): ₹129,551 (constant INR)(23)
- Employment-to-Population Ratio (EPR) for adults: 53·31% (24)
- One US Dollar (USD) = Indian Rupees (INR) 83·67 in 2024(25)
- One International Dollar (Int$) = INR 20·4 in 2024(26)

### Model Parameter Estimations

The economic burden was estimated across three pathways, with full equations in the appendix.

DHL leads to a lower likelihood of employment; literature suggests a 12·5%-16·5% lower chance of employment for individuals with HL compared to those without HL. (4) DL was estimated by multiplying the number of unemployed adults (18-60 years) with the per-capita GNI due to labor force.

Indirect Loss of GNI (Caregiver Productivity Loss), and due to the absence of HL-specific data, caregiver productivity loss was conservatively assumed at 10%, extrapolated from blindness studies and held constant across age groups.(20) IL is modelled for different age groups and HL severity, assuming that all children with HL need caregiving and only adults with DHL require caregiving. The total IL is the sum of all the loss across these age groups and HL severity. Adult DHL prevalence for the 18–60 and 60+ bands is derived from age-stratified estimates,(27) population-weighted using Census 2011 age-structure proportions scaled to 2024 population totals.(28)

For the Loss of GNI due to Low Education, hearing loss (HL) is associated with a lower probability of educational attainment, which leads to a unemployment or underemployment in the adulthood. We used the Population Attributable Fraction (PAF)(29) method to convert a baseline outcome probability of low educational attainment (in the non-HL population) into an HL-specific probability, then compute the excess number of cases with education loss attributable to HL.

### Sensitivity Analysis

We performed sensitivity analyses on the estimated parameters. Adhering to professional guidelines on cost-of-illness studies and health economics, we have conducted One-way Sensitivity Analysis (OWSA) and Probabilistic Sensitivity Analysis (PSA) on the parameters,(30) a summary of which is mentioned in the Table 2 Table 2.

**Table 2:** Summary of Type of Sensitivity Analyses on each category.

| Pathway | Target Population | Outputs | Sensitivity Analysis |
| --- | --- | --- | --- |
| 1. Direct GNI Loss (DL) | Adults 18–60 with DHL | Lost income from non-employment | OWSA + PSA |
| 2. Indirect GNI Loss (IL) | Children (<18) with HL, Adults 18+ with DHL | Caregiver productivity loss | OWSA + PSA |
| 3. Cost of Low Education (LE) | Adults with HL (18+) | Downstream impact of poor education on adult income | OWSA + PSA |

OWSA was conducted by varying individual parameters within plausible bounds to assess their influence on each cost pathway. Key parameters and their ranges are shown in Table 3.

**Table 3:**
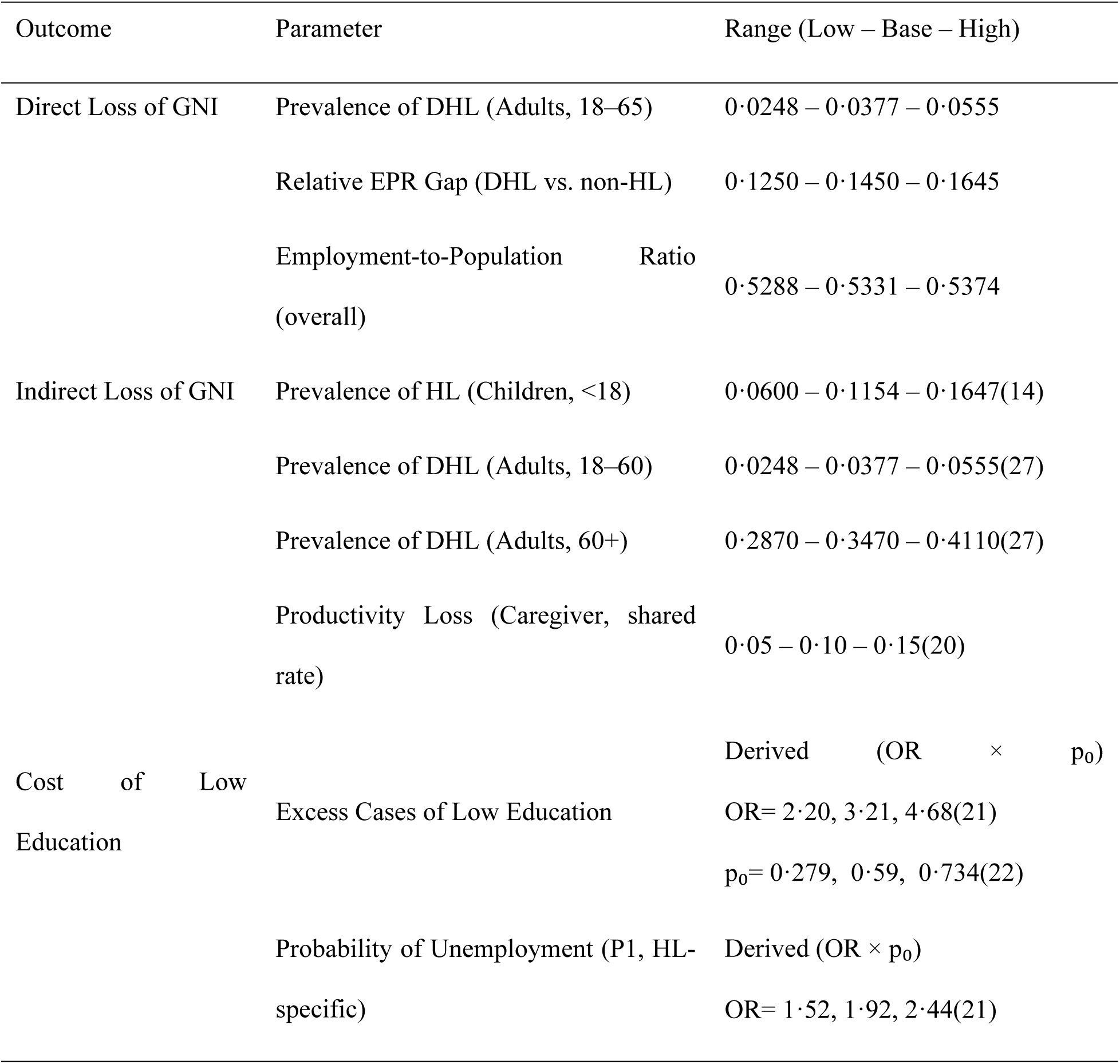
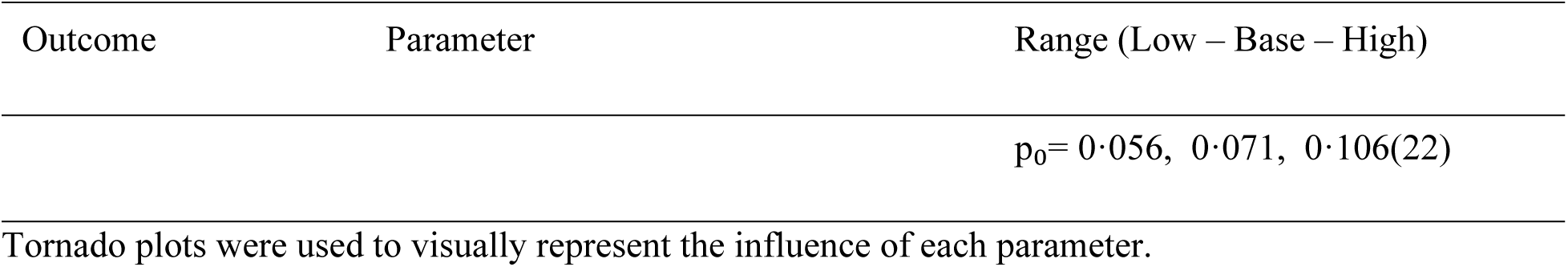
OWSA Parameters.

**Table 4:** Summary of Economic Burden by Cost Pathway (2024, India)

All figures in Billion INR, with Billion USD in parentheses (1 USD = 83·67 INR)
| Cost Component | Billion INR (Billion USD) |  |  |
| --- | --- | --- | --- |
|  | Mean | Median | IQR |
| Direct Loss (DL) | 4,648·4 (55·6) | 4,592·0 (54·9) | 1,183·6 (14·1) |
| Indirect Loss (IL) | 3,296·1 (39·4) | 3,210·9 (38·4) | 1,185·0 (14·2) |
| Total Economic Burden of HL | 7,944·5 (94·6) | 7,802·9 (93·3) | 2,368·6 (28·3) |
| Cost of Low Education (LE)* | 1,229·5 (14·7) | 1,108·0 (13·2) | 751·6 (9·0) |

\* The low education pathway was not summed with Direct Loss because this would partially double-count productivity losses arising through unemployment.

PSA assessed combined uncertainty across all pathways using beta distributions for parameters such as prevalence, EPR, and GNI loss proportions. A beta distribution is used for proportions and probabilities conforming to guidelines in health economic modelling.(31)

The number of excess cases of low educational attainment attributable to hearing loss - a non-negative, right-skewed count variable derived from the odds-ratio-based excess-case calculation, was modeled using a Gamma distribution

Monte Carlo simulations (10,000 iterations) generated cumulative cost distributions, with key percentiles (2·5th, 50th, 97·5th) reported. Results were plotted using cumulative distribution functions (CDFs). Distribution parameters were estimated using the method of moments; the fitting procedure and its rationale are detailed in the appendix.

## Results

### Total Number of People with HL and DHL in India

The data was stratified into three age groups; children (<18 years), adults (18-60 years), and elderly (>60 years). The prevalence of HL and DHL and the total number of people with HL and DHL in each age group are presented in Table 1.

Three Economic Loss Pathways

The economic loss due to HL and DHL are estimated in three pathways.

### Direct Loss of GNI (DL)

The median (IQR) total income lost due to unemployment among working-age adults with DHL is INR 4,354·65 billion (USD 52 billion). The OWSA variation was INR 2,870 – INR 6,397, depending most sensitively on prevalence of DHL. PSA yielded a median loss of INR 4,592 billion (USD 54·9) and IQR INR 1,183·6 (USD 14·1). The Tornado plot in Figure 2 illustrates OWSA results, and the Cumulative Density Function(CDF) plot in Figure 3 illustrates the PSA results for loss of employment due to DHL.

**Figure 2:**
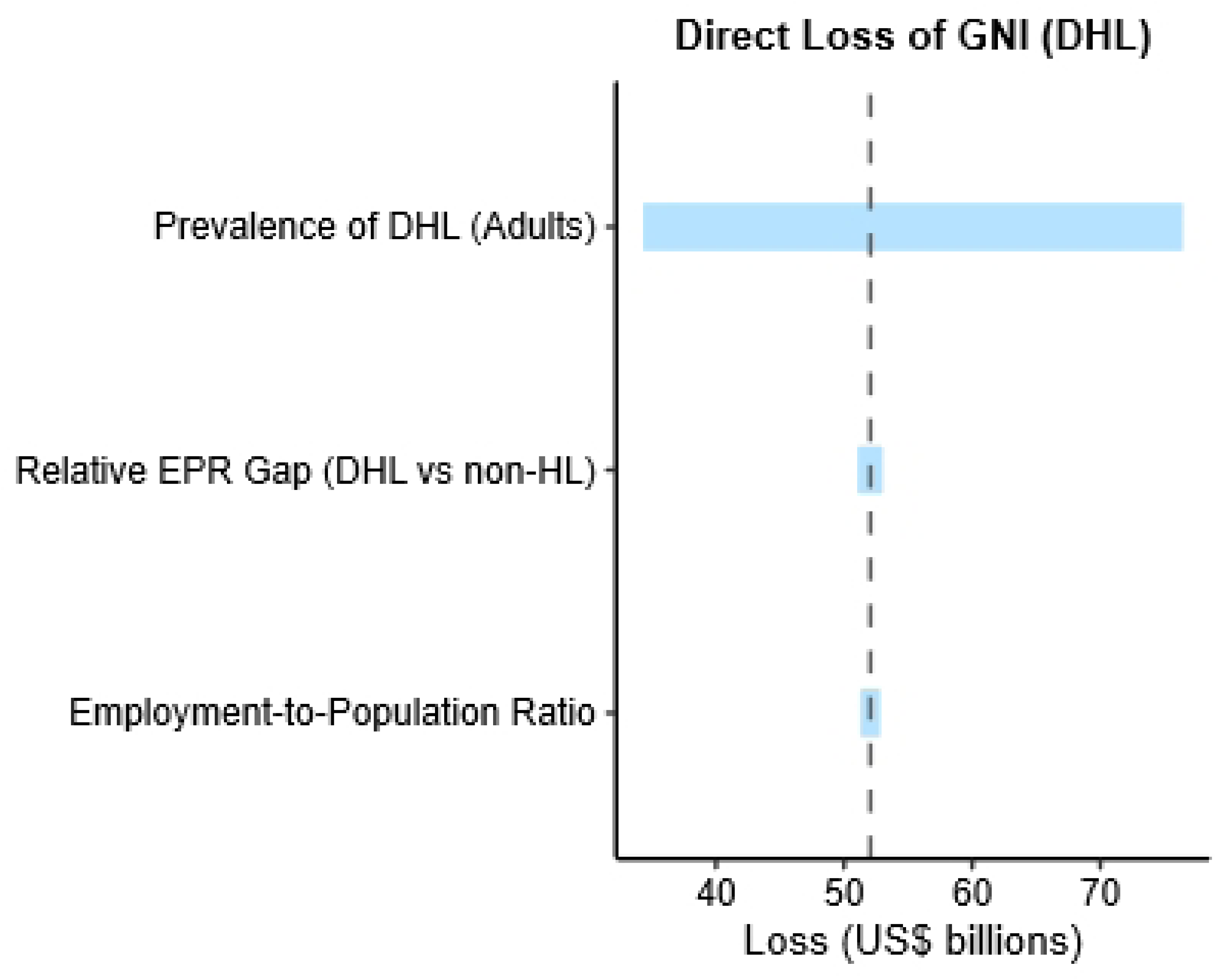
Tornado Plot of OWSA – DL due to DHL

**Figure 3:**
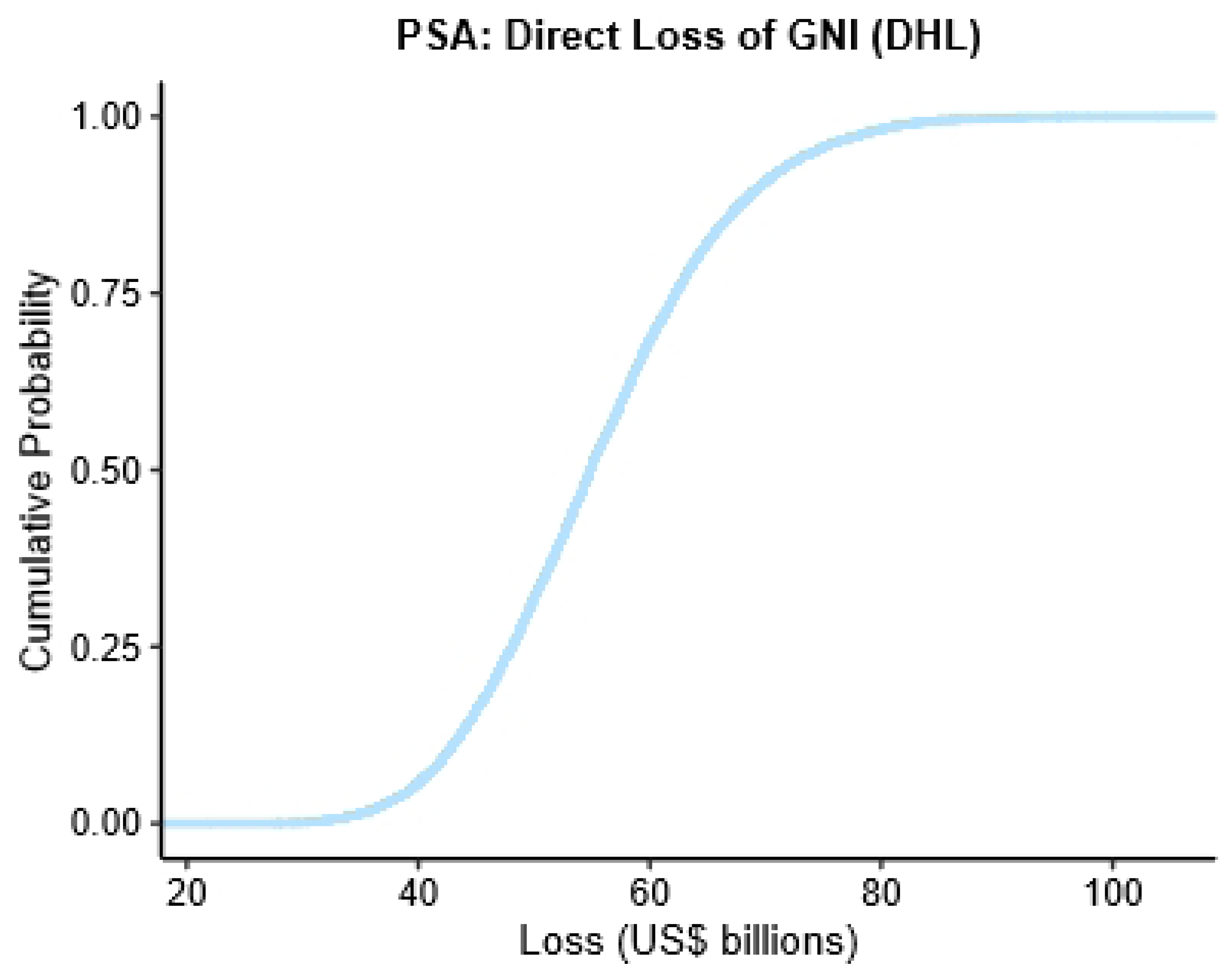
CDF plot of PSA – DL due to DHL

### Indirect Loss of GNI (IL) – Caregiver economic burden

The median (IQR) total cost of caregiver burden due to DHL is INR 3,268 billion (USD 39 billion). The OWSA showed the loss varied between INR 1,634 billion (USD 19·5 billion) and INR 4,902 billion (USD 41·9 billion), depending most sensitively on the prevalence of DHL amongst those aged 18 to 60 years. The results of OWSA are presented in the tornado plot in Figure 4 Figure 3. PSA yielded a median loss of INR 3,210·9 billion (USD 38·4 billion) and IQR of INR 1,185 billion (USD 14·2 billion). The Cumulative Density Function plot in Figure 5 illustrates the PSA results for IL.

**Figure 4:**
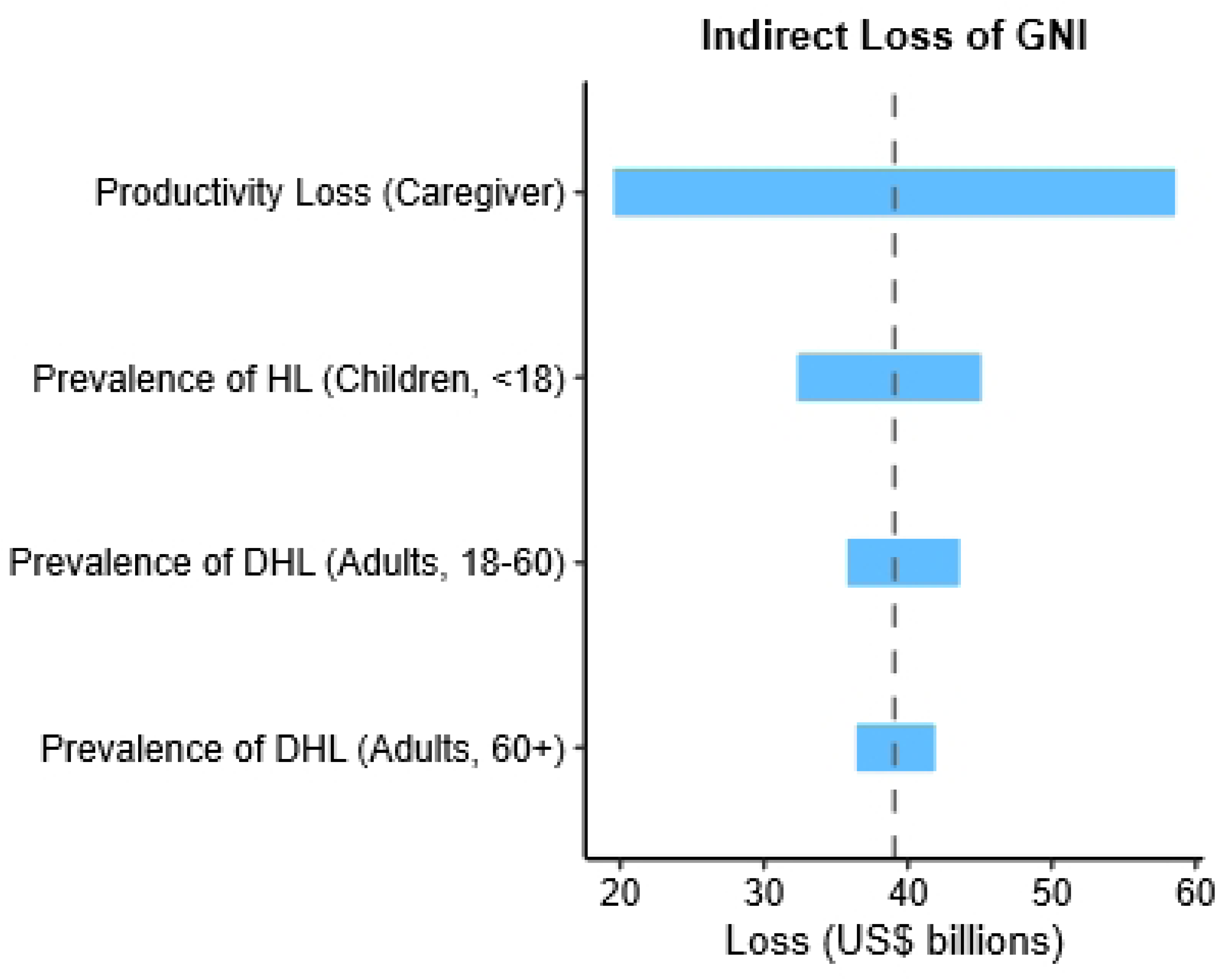
Tornado Plot: IL due to HL and DHL

**Figure 5:**
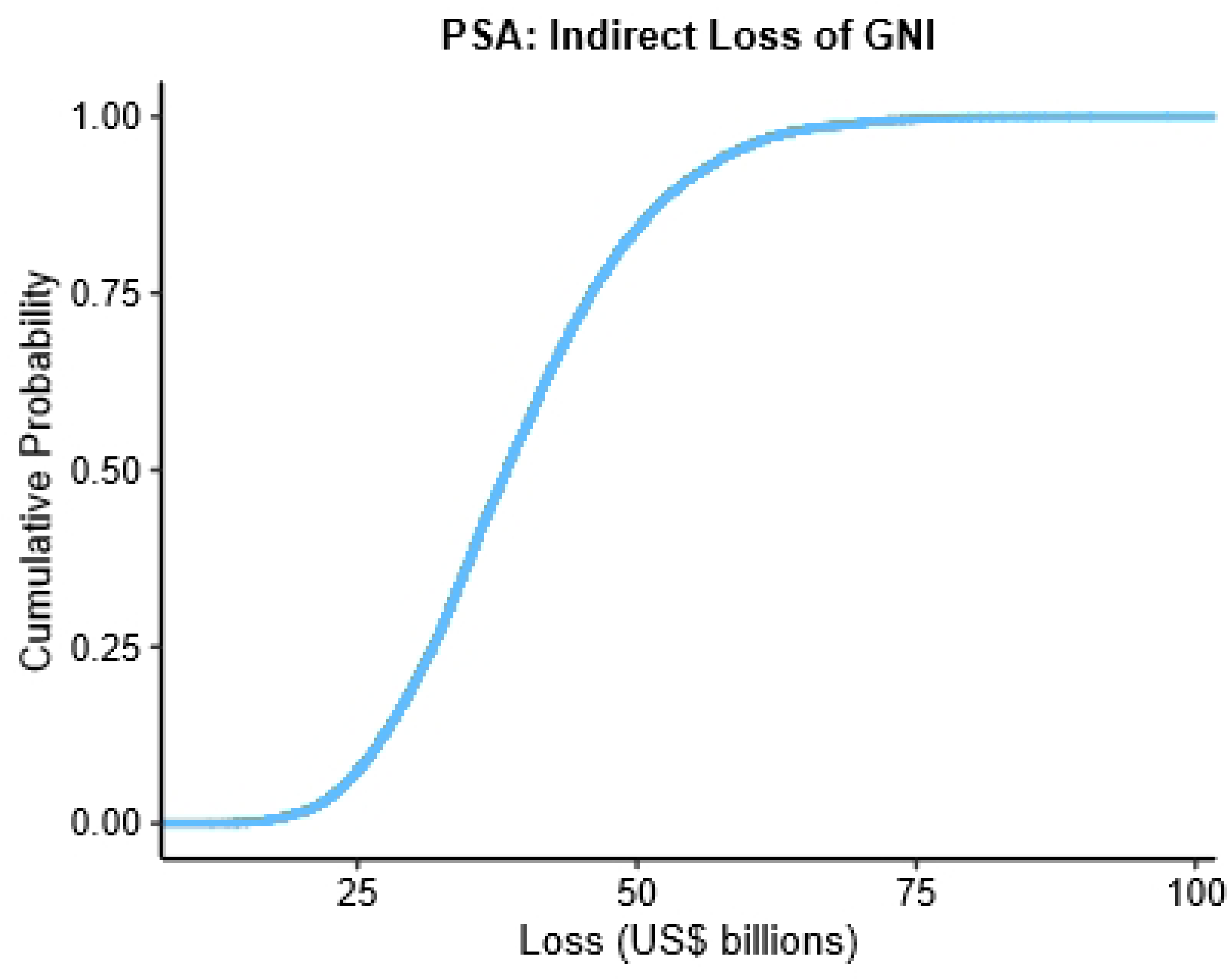
CDF plot of Cost of Caregiver Burden due to DHL

### Loss of GNI due to Low Education (LE)

The total excess number of people with low education attributed to HL are 33,506,880. The median cost of low education is INR 1,041·9 billion (USD 12·45 billion); OWSA varied between INR 559 billion (USD 6·6 billion) to INR 1,827 billion (USD 21·8 billion), the tornado plot in illustrates the results of each parameter variation. The PSA resulted in a median of INR 995·5 billion (USD 11·9 billion) and IQR of INR 648·6 billion (USD 7·7 billion). The tornado plot in Figure 6 illustrates the results of OWSA and the Cumulative Density Function plot in Figure 7 illustrates the PSA results for the cost of low education.

**Figure 6:**
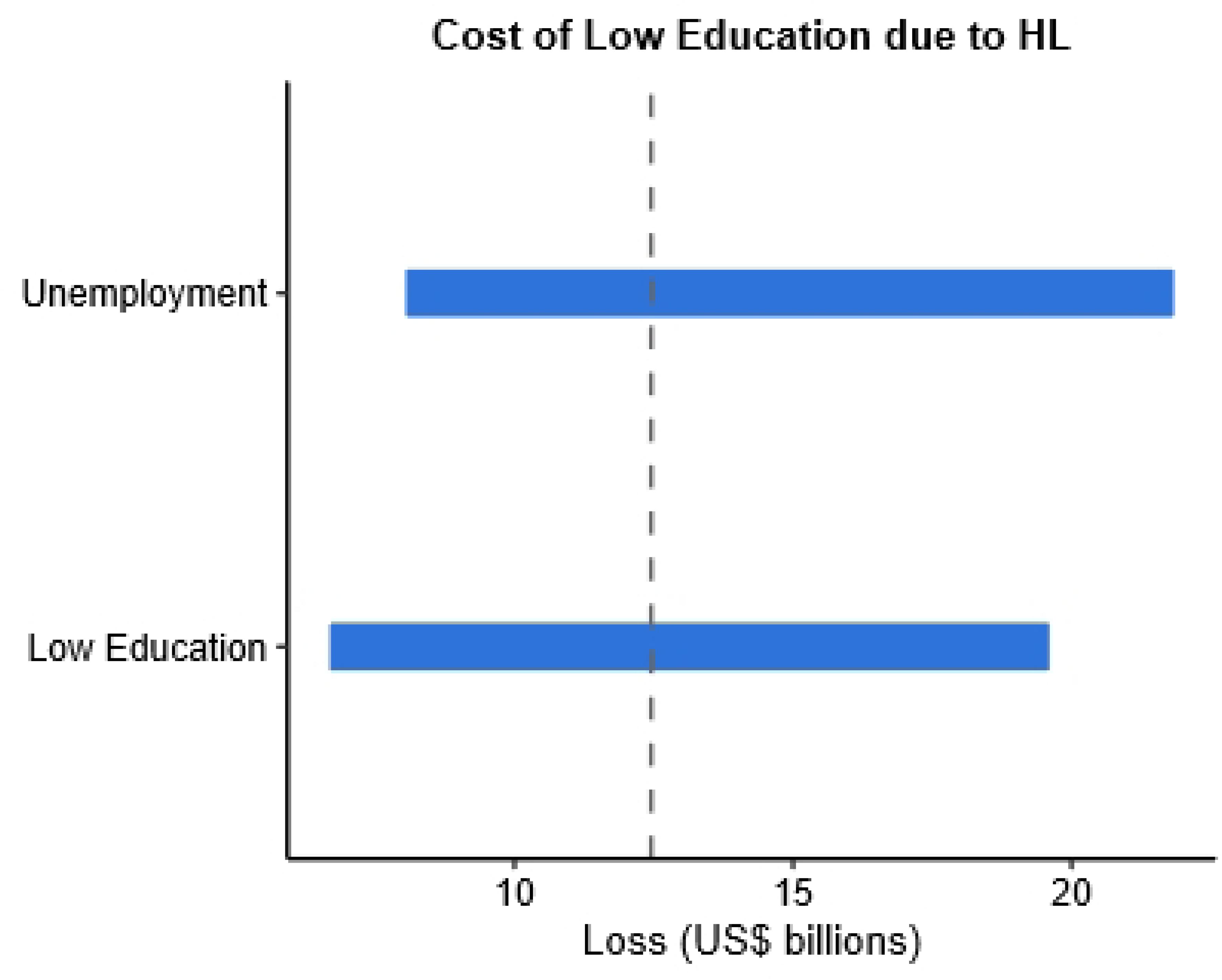
OWSA: Loss of GNI due to LE amongst those with HL

**Figure 7:**
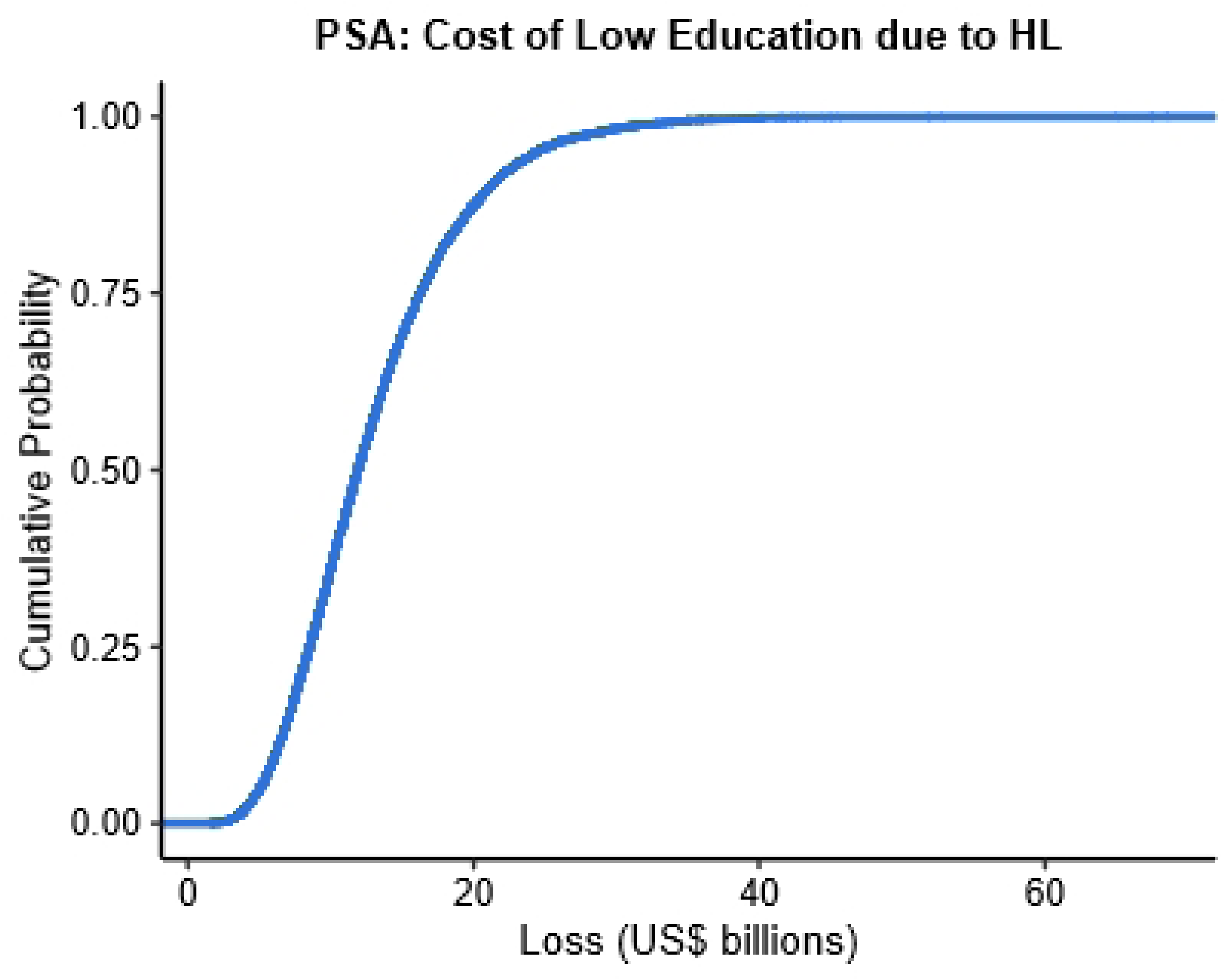
CDF plot of Cost of Low Education Attainment amongst Children with HL

## Discussion

To our knowledge, this is among the first India-focused societal cost-of-illness (COI) studies estimating the economic burden of hearing loss (HL) using nationally derived demographic and macroeconomic parameters. The study demonstrates that the economic burden of HL in India is substantial and multidimensional, extending beyond productivity losses alone. This study estimates the economic burden of HL across three different pathways (productivity loss due to unemployment, lost productivity of the caregiver, and lost income due to low education). Estimating the economic burden across different pathways has been undertaken before in relation to visual impairment, and it presents the multi-dimensional socio-economic impact of a health condition.(32)

### Three Economic Pathways

We have considered unemployment due to Disabling Hearing Loss (DHL) for the estimation of the direct loss of GNI (DL) as this is considered to impact an individual’s ability to work. Due to a lack of wide literature on the prevalence of DHL amongst adults in India, we have relied on a very few studies for our estimations, hence we have observed wide range in the DL estimation. However, the OWSA and PSA have accounted for the variations in the parameters. According to the Census 2011 data, only about 26% of those with disabilities and in working age group were employed,(28) our estimation has considered this to be 46·7% as per WHO report, leading to a more conservative estimation of DL in India.(14)

The Indirect Loss of GNI (IL) due to HL was estimated based on age stratification and caregiver burden, where the age stratified analysis was based on the assumption that all children with HL and adults with DHL would require some caregiving, we have considered that caregivers spend 10% of their time caring for children and adults with HL and DHL respectively. Although there are studies understanding various aspects of caregiving such as psychological wellbeing(33,34) there is a need for quantifying the time lost by the caregivers caring for individuals with HL and DHL. The age stratification allows us to estimate the IL at a more granular level. We have observed that HL amongst children leads to highest variation after the caregiver burden which was varied between 5% and 15%, this aligns with finding from literature that children would require greater amount of caregiving(33).

Our estimation of the Loss of GNI due to Low Education was based on the assumption that as a consequence of low education, there would be unemployment amongst those with HL. Since we could not find any studies linking HL and education attainment in India, we have relied on estimations from a US study on the odds of having low education amongst those with and without HL and odds of being unemployment due to low education. (21) The proportion of individuals with HL and Low Education and Unemployment was derived using the Population Attributable Factor (PAF) method which is used for attributing a risk to an exposure such as low education and HL in our study. (35) We observed a wide range along with a right skewed estimation which is due to the odds more than doubling from low to high for low education in the US study.(21) Although the adjusted odds ratio was used to account for all the confounders for low education amongst those with HL, the estimation has to be considered with caution because of the difference in education system and the socio-economic conditions related to India. The economic burden (loss of GNI due to low education) quantified in this study is, in principle, reducible through interventions which are already proven to be cost-effective in India such as deaf education and cochlear implants.(13,36)

This study presents a DL of Int$ 225 billion (USD 54·9 billion) which is substantially higher than the previous estimates of Int$ 9·6 billion reported by the World Health Organization.(14) However, these two estimates differ methodologically, firstly, our study estimated the cost based on those with DHL whereas the WHO study estimated based on moderately severe HL; secondly, our estimations were based on India specific data related prevalence and productivity parameters, and the WHO estimation was based primarily on extrapolated assumptions from the UK and Australia.

The study highlights the need for India specific data related to various aspects of HL such as HL attributable low education, disaggregated prevalence estimations, and DHL estimations. Since India is a diverse country with 29 states and union territories, the socio-economics and the health statuses change in each state and region, hence there is a need for greater number of disaggregated studies to inform studies such as COI analyses and for disability inclusive planning at national level.(37,38)

### Strengths and Limitations

A major strength of this study is that we have estimated the economic burden across three major pathways in a macro-economic societal perspective stratified for different age groups. We have used both OWSA and PSA to account for the various parameter uncertainties in the data and thereby making the model robust under various assumptions. We have estimated the loss of GNI due to low education (LE), an area often missed in COI analyses due to lack of underlying data that attribute HL and education respectively. We too could not find India specific data and used the PAF approach to derive the LE estimates based on the odds ratio from a US study which limits the generalizability of this estimate albeit presents a method for economic burden estimation due to low education attributable to a risk factor such as HL. Due to the lack of 2024 population data, we relied on extrapolating the age-wise population based on age proportions from the 2011 census data, however, the sensitivity analyses account for these variations in the data. The wide range presented in the sensitivity analysis of DL, IL, and LE should be interpreted as a reflection of the underlying uncertainty related to the epidemiological evidence, the socio-economic parameters and risk parameters. There is a need for caregiver burden estimation related to HL as we have used proxy estimation due to the lack of HL specific data. The burden estimated in this study is likely to fall disproportionately on children, older adults, and households providing informal care. Future studies should evaluate geographic, socioeconomic, and gender-related inequalities in hearing-loss-associated economic burden.

### Policy Implications

The findings of this study have important implications for research, policy, financing, and advocacy related to hearing loss in India. The age stratified analysis helps in prioritizing already established cost-effective(39) screening and timely interventions according to the age groups.

Strengthening rehabilitation pathways may improve labor force participation and reduce productivity losses attributable to DHL.

## Data Availability

All the data sources and data are already listed in the manuscript and supplementary material.

## Author Contributions

SM and VR conceived this study. SM prepared the data, led the analysis, and wrote the first draft of the manuscript. VR provided feedback during the design, analysis, and interpretation of the study. VR and GVS contributed to the revisions of the manuscript. All authors had access to the data and analysis and have contributed to reviewing and editing of the manuscript and approved of the final text. All authors made the final decision to submit the article for publication.

## Data Sharing

The data sources used in this study and the specific analyses datasets are available upon request to the corresponding author.

## Acknowledgements

Authors acknowledge the support from Sri Ramachandra Institute of Higher Education and Research (SRIHER) in publishing this article.

## Supporting Information

The S1 supporting information document has supporting information about the parameter estimation methods and also sensitivity analyses.

## References

1 World report on hearing. Geneva: World Health Organization, 2021.

2 World Health Organization. State of Hearing and Ear Care: Regional Reports and Analyses. 2014.

3 United Nations D of E and SAPD. World Population Prospects 2024. 2024 https://population.un.org/wpp/ (accessed July 1, 2026).

4 World Health Organization. Global costs of unaddressed hearing loss and cost-effectiveness of interventions: a WHO report, 2017. Geneva: World Health Organization, 2017.

5 Kim SY, Min C, Yoo DM, et al. Hearing Impairment Increases Economic Inequality. Clin Exp Otorhinolaryngol 2021; 14: 278–86.

6 Leverton T. Depression in older adults: hearing loss is an important factor. BMJ 2019; : l160.

7 Gupta S, Jaiswal A, Sukhai M, Wittich W. Hearing disability and employment: a population-based analysis using the 2017 Canadian survey on disability. Disabil Rehabil 2023; 45: 1836–46.

8 Neitzel RL, Swinburn TK, Hammer MS, Eisenberg D. Economic impact of hearing loss and reduction of noise-induced hearing loss in the United States. Journal of Speech, Language, and Hearing Research 2017; 60: 182–9.

9 Johns B, Baltussen R, Hutubessy R. Programme costs in the economic evaluation of health interventions. Cost Effectiveness and Resource Allocation 2003; 1: 1–10.

10 Ramkumar V, John KR, Selvakumar K, Vanaja CS, Nagarajan R, Hall JW. Cost and outcome of a community-based paediatric hearing screening programme in rural India with application of tele-audiology for follow-up diagnostic hearing assessment. Int J Audiol 2018; 57: 407–14.

11 Sahoo KC, Dwivedi R, Athe R, et al. Cost-effectiveness of portable-automated ABR for universal neonatal hearing screening in India. Front Public Health 2024; 12: 1364226.

12 Sahoo RK, Sahoo KC, Pattanayak U, et al. Economic evaluation of hearing aid use and quality of life in older adults with hearing impairment in India. Front Med Technol 2026; 8: 1800134.

13 Sharma A, Prinja S, Thakur R, et al. Healthcare Cost of Cochlear Implantation in India. Indian Journal of Otolaryngology and Head & Neck Surgery 2023; 76: 1716.

14 Global costs of unaddressed hearing loss and cost-effectiveness of interventions A WHO Report, 2017. 2017.

15 Inciong JFB, Chaudhary A, Hsu HS, et al. Economic burden of hospital malnutrition: A cost-of-illness model. Clin Nutr ESPEN 2022; 48: 342–50.

16 Kondapura MB, Manjunatha N, Nagaraj AKM, et al. Cost of Illness Analysis of Common Mental Disorders: A Study from an Indian Academic Tertiary Care Hospital. Indian J Psychol Med 2023; 45: 519–25.

17 Borre ED, Diab MM, Ayer A, et al. Evidence gaps in economic analyses of hearing healthcare: A systematic review. EClinicalMedicine 2021; 35: 100872.

18 International Labour Organization, World Bank, Our World in Data. Employment Rate – ILO. Our World in Data, 2026 https://ourworldindata.org/grapher/employment-to-population-ratio.

19 The World Bank. World Development Indicators. The World Bank Group. 2019. https://data.worldbank.org/indicator/SP.POP.TOTL?locations=IN (accessed Jan 7, 2025).

20 Mannava S, Borah R, Shamanna B. Current estimates of the economic burden of blindness and visual impairment in India: A cost of illness study. Indian J Ophthalmol 2022; 70: 2141.

21 Emmett SD, Francis HW. The socioeconomic impact of hearing loss in U.S. adults. Otology and Neurotology 2015; 36: 545–50.

22 Ministry of Statistics and Programme Implementation. Periodic Labour Force Survey (PLFS) Annual Report JULY 2023 - JUNE 2024. New Delhi, 2024.

23 World Bank. GDP Per Capita (Constant LCU) – India. 2026. https://data.worldbank.org/indicator/NY.GDP.PCAP.KN?locations=IN.

24 International Labour Organization. Employment to population ratio, 15+, total (%) (national estimate) - India. 2025.

25 World Bank. Official Exchange Rate (LCU per US$, Period Average) – India. 2026. https://data.worldbank.org/indicator/PA.NUS.FCRF?locations=IN.

26 International Monetary Fund. Implied PPP Conversion Rate – India. 2026. https://www.imf.org/external/datamapper/PPPEX@WEO/OEMDC/ADVEC/WEOWORLD/IND.

27 T B, I M, H K, GV M, S P. Prevalence of hearing impairment in Mahabubnagar District, Telangana State, India. Ear Hear 2019; 40: 204–12.

28 Census of India. Office of the Registrar General & Census Commissioner. 2011. http://censusindia.gov.in/.

29 Mansournia MA, Altman DG. Population attributable fraction. BMJ 2018; : k757.

30 Jo C. Cost-of-illness studies: concepts, scopes, and methods. Clin Mol Hepatol 2014; 20: 327.

31 Briggs AH, Weinstein MC, Fenwick EAL, Karnon J, Sculpher MJ, Paltiel AD. Model Parameter Estimation and Uncertainty Analysis. Medical Decision Making 2012; 32: 722–32.

32 Wong B, Singh K, Khanna RK, et al. The Economic and Social Cost of Visual Impairment and Blindness in India. California, USA, 2021 https://www.seva.org/site/DocServer/Seva_Cost_of_Visual_Impairment_in_India.pdf (accessed Sept 10, 2022).

33 Kumar N, Samantray VR, Syed KA, Magendiran N, John M. Psychological and Economic Burden on Caregivers in State-funded Pediatric Cochlear Implantation in South India – A Pilot Study. 2026; 24: 135–40.

34 Van Driessche A, Jotheeswaran AT, Murthy GVS, et al. Psychological well-being of parents and family caregivers of children with hearing impairment in south India: Influence of behavioural problems in children and social support. International Review of Psychiatry 2014; 26: 500–7.

35 Haresabadi M, Esfahani EN, Mansournia MA, et al. Model-based estimation of the generalized impact fraction (GIF) and population attributable fraction (PAF) for interventions targeting risk factors affecting the five-year risk of diabetic retinopathy. BMC Public Health 2025; 25: 2667.

36 Emmett SD, Sudoko CK, Tucci DL, et al. Expanding Access: Cost-effectiveness of Cochlear Implantation and Deaf Education in Asia. Otolaryngology–Head and Neck Surgery 2019; 161: 672– 82.

37 Gvs M, Velarde RM. The Disability Data Report 2023 Fordham Research Consortium on Disability ACKNOWLEDGEMENTS. 2023. http://www.ace.disabilitydata.fordham.edu (accessed July 26, 2026).

38 Haile LM, Kamenov K, Briant PS, et al. Hearing loss prevalence and years lived with disability, 1990-2019: Findings from the Global Burden of Disease Study 2019. The Lancet 2021; 397: 996–1009.

39 Tordrup D, Smith R, Kamenov K, Bertram MY, Green N, Chadha S. Global return on investment and cost-effectiveness of WHO’s HEAR interventions for hearing loss: a modelling study. Lancet Glob Health 2022; 10: e52–e62.

